# On period, cohort and population life expectancy

**DOI:** 10.64898/2026.03.16.26348495

**Authors:** Djuly Asumpta Pierre-Paul, Valentin Rousson, Isabella Locatelli

## Abstract

Period Life Expectancy (PLE) is a measure of longevity valued for its sensitivity to short and long-term changes. However, it refers to a hypothetical cohort, not to a real population, thereby undervaluing longevity under declining mortality conditions. Other measures such as the Average Cohort Life Expectancy (ACLE) only partially overcome this limitation, still underestimating population longevity. This article introduces a new indicator, the Population Life Expectancy (PoLE), defined as the mean age at death of active cohorts in the studied population. Using a log-linear Poisson model with age-period interaction to project mortality of non-extinct cohorts, we estimated PoLE in Switzerland and Norway over 1876-2024, and compared it to PLE, Cohort Life Expectancy (CLE), and ACLE. PoLE clearly exceeded PLE, increasing from 63.3 to 89.7 for Swiss men (PLE from 37.7 to 82.4), and from 65.4 to 91.3 for Swiss women (PLE from 41.4 to 85.9), revealing a gain of about +50% over 150 years, rather than +100% suggested by PLE. Comparable results were obtained in Norway. PoLE was also higher than CLE until the mid-20th century, when the relation reversed, indicating that life expectancy is now higher for newborns than for those already alive, a tangible sign of human progress.

## Introduction

Life expectancy is probably the best known and most widely used indicator of longevity [1]. A distinction is made between the concepts of *Cohort* life expectancy (CLE) and *Period* life expectancy (PLE) for a given year. The former is defined as the average age at death of the cohort of individuals born in that year, the latter as the average age at death of a hypothetical cohort of individuals who would live their entire lives under the mortality conditions of the year. These are also referred to as CLE or PLE *at birth*. One can similarly define a (remaining) CLE or PLE *at a given age* as the average number of years remaining to be lived by the individuals of the observed, respectively hypothetical, cohort who are still alive at that age. By definition, the CLE can only be calculated for cohorts that are extinct or almost extinct. For example, for Switzerland and Norway, the two countries considered in this study, CLE is only available for cohorts up to 1933 in the Human Mortality Database (HMD), a freely accessible demographic database. In contrast, PLE can be calculated until recent years (up to 2024 for both countries in the HMD), which is a considerable advantage.

Another reason explaining the popularity of the PLE indicator is that it is highly sensitive to both acute health crises and long-term societal trends. For example, in 1918 during the Spanish flu, PLE fell by around ten years in Switzerland and Norway (while CLE was barely affected that year). Another example is that of East-Germany, where PLE gradually caught up that of West Germany after the reunification in 1989 [2]. More recently, the PLE demonstrated a stagnation or even a fall in the US attributed to the crisis of obesity epidemic and over-consumption of medication [3]. During the COVID-19 pandemic, this indicator has also been widely used to quantify the impact of the sanitary crisis on mortality [4-6]. For example, in Switzerland, PLE fell by around 8 months in 2020, before returning to its pre-pandemic level in 2021, then falling again by 2-3 months in 2022 due to the summer heatwave [7-9]. In Norway, PLE was not impacted in 2020-21 [4], but it decreased by more than 6 months in 2022 (HMD).

On the other hand, PLE has often been misinterpreted [1,10]. One major difficulty is that this indicator refers to a hypothetical cohort rather than to a real population. In fact, a real population, e.g. all people living in Switzerland or Norway in a given year, has not lived and will not live its entire life during that year. Rather, a population consists of a mixture of “active cohorts” of individuals born in different years in the past and likely to continue living in the future. If mortality is expected to decline in the future (according to past trends), the PLE calculated that year will underestimate the average age at death of this real population, and even more so if a catastrophic event, such as the Spanish flu, occurs that year. Recognizing the inability of PLE to represent the life expectancy of such a real population, demographers have been attempting since the 1980s to develop other longevity indicators.

A first attempt was made by Nicolas Brouard, who proposed in a French journal the idea of a “durée de vie moyenne actuelle compte tenu de l’histoire passée de la mortalité” [11]. This concept was then updated and formalized several years later into what is known as the *Cross-sectional Average Length of life* (CAL) [12]. This indicator is defined as the sum of the proportions of survivors among active cohorts in a given year, thus only considering the *past* mortality experience of these cohorts, not the *future*. As a result, the CAL indicator will systematically be lower than PLE in a context of declining mortality, which paradoxically suggests that PLE would overestimate the longevity of this real population. This inconsistency has been highlighted by several demographers, such as Goldstein and Watcher [13,14].

Another significant attempt was made by Schoen and Canudas-Romo in 2005 with the introduction of the *Average Cohort Life Expectancy* (ACLE) [15]. This indicator differs from CAL in that it considers past *and* future mortality of the active cohorts in a given year, which requires some mortality projections. The ACLE is defined as a weighted average of the life expectancies at birth of all these cohorts, assuming for non-extinct cohorts a future annual mortality decline of 0.5% at each age. Contrary to CAL, the ACLE indicator potentially exceeds the PLE in such a favorable mortality context, which is a desirable property. However, it still considerably underestimates the intended “average length of life of cohorts alive at a given time”, as we shall see below. Also, the assumption of an age-independent mortality decline is not truly consistent with observed mortality patterns.

Until very recently (as late as 2021) still other proposals have been made. Some indicators, such as the *Tempo-Adjusted Life Expectancy* (TALE) and the *Lagged Cohort Life Expectancy* (LCLE), follow a similar approach to CAL, and all three measures have been shown to approximate each other under quite general mortality conditions [16-17]. Other attempts consisted of derivatives or extensions of CAL, such as the *Truncated* CAL (TCAL), the *Childless* CAL (CALL), and the CAL *for inequality* (CAL+) [18-20].

In this article, we introduce a novel indicator, which we called *Population Life Expectancy* (PoLE), designed to more directly answer the above question raised by demographers of how long a real population of individuals alive in a given year can expect to live. To achieve this, a simple yet robust prediction model is used to extrapolate future mortality trends and life expectancies of active cohorts. This new indicator is calculated and compared with PLE, CLE, and the ACLE indicators, using HMD data from Switzerland and Norway.

## Methods

We introduce here a novel life expectancy indicator, the *Population Life Expectancy* (PoLE), defined as the mean age at death of the population alive a given year *y* ≤ *y* ^*\**^, where *y* ^*^ is the last year with available data. This new indicator is inspired by the A*verage Cohort Life Expectancy* (ACLE) proposed by [15]. The latter was defined as the average life expectancy *at birth* of all cohorts active in year *y*, weighted by the percentage of each cohort still alive that year, where mortality projections needed for life expectancies of non-extinct cohorts were made assuming an annual decline in mortality of *c* = 0.5% at each age (eq. (6a) in [15]).

The PoLE indicator introduced in this paper modifies the ACLE in three ways. It is obtained by averaging the *expected age at death* for individuals at each age *a* in year y, 0 ≤ *a* ≤ 110, instead of their life expectancies at birth (first difference), weighting by the population age structure of year *y*, instead of by the percentage of the corresponding cohorts still alive in this year (second difference). Considering that the expected age at death of an individual of age *a* in year *y* can be obtained by adding this age (already reached) to the *remaining* life expectancy at age *a* of the cohort born *a* years earlier (in the year *y* − *a*), the new indicator will take the form:

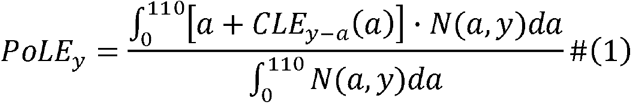

Here, *N* (*a,y*)is the size of population of age *a* in year *y*, and *CLE*_*y-a*_*(a)* is the remaining cohort life expectancy at age *a* for the cohort born in year *y* − *a*. The last can be written as follows:

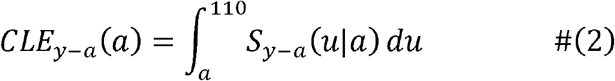

where *S*_*y-a*_ *(u*|*a*) is the conditional survival function defining the probability of surviving up to age *u* (*a* ≤ *u* ≤ 110) for an individual of the cohort *y* − *a* being still alive at age *a*:

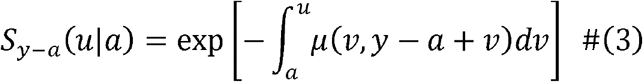

In equation (3), the quantity *μ* (*v,y* − *a* +*v*) represents the *force of mortality* at age *v* in year *t* = *y* - *a* + *v*, with 0 ≤ *a* ≤ 110, *v* ≥, and *y* ≤ *y*^*^. Empirical estimates of *μ* (*v, t*) are obtained dividing the number of deaths at age *v* in year *t* by the mid-population at the same age between January 1^st^ and December 31 of the same year.

Since not all cohorts involved in (3) are extinct, only a part of the *μ*’s needed to derive the PoLE for a given year *y* can be empirically estimated, namely those for which *t* = *y* − *a* + *v* ≤ *y*^*^, meaning that, for an individual of age *a* in year *y* (belonging to the cohort *y* − *a*), mortality should be projected for ages *v* > *y*^*^− *y* + *a*. For example, if the last year with observed data is *y*^*^ = 2024, and we want to estimate the PoLE for *y* = 1990, mortality of individuals of age 80 will be entirely known until the extinction of the cohort (they will be 110 years old in 2020 ≤ *y* ^*^), while mortality of individuals aged 30 (born in 1960) should be projected from age 2024 − 1990+ 30 = 64 until age 110, i.e. until year 2070 (Figure 1). To obtain such mortality projections, we relied on a projection model, which represents the third difference with respect to [15] who assumed a 0.5% annual decline in mortality at each age.

**Figure 1.**
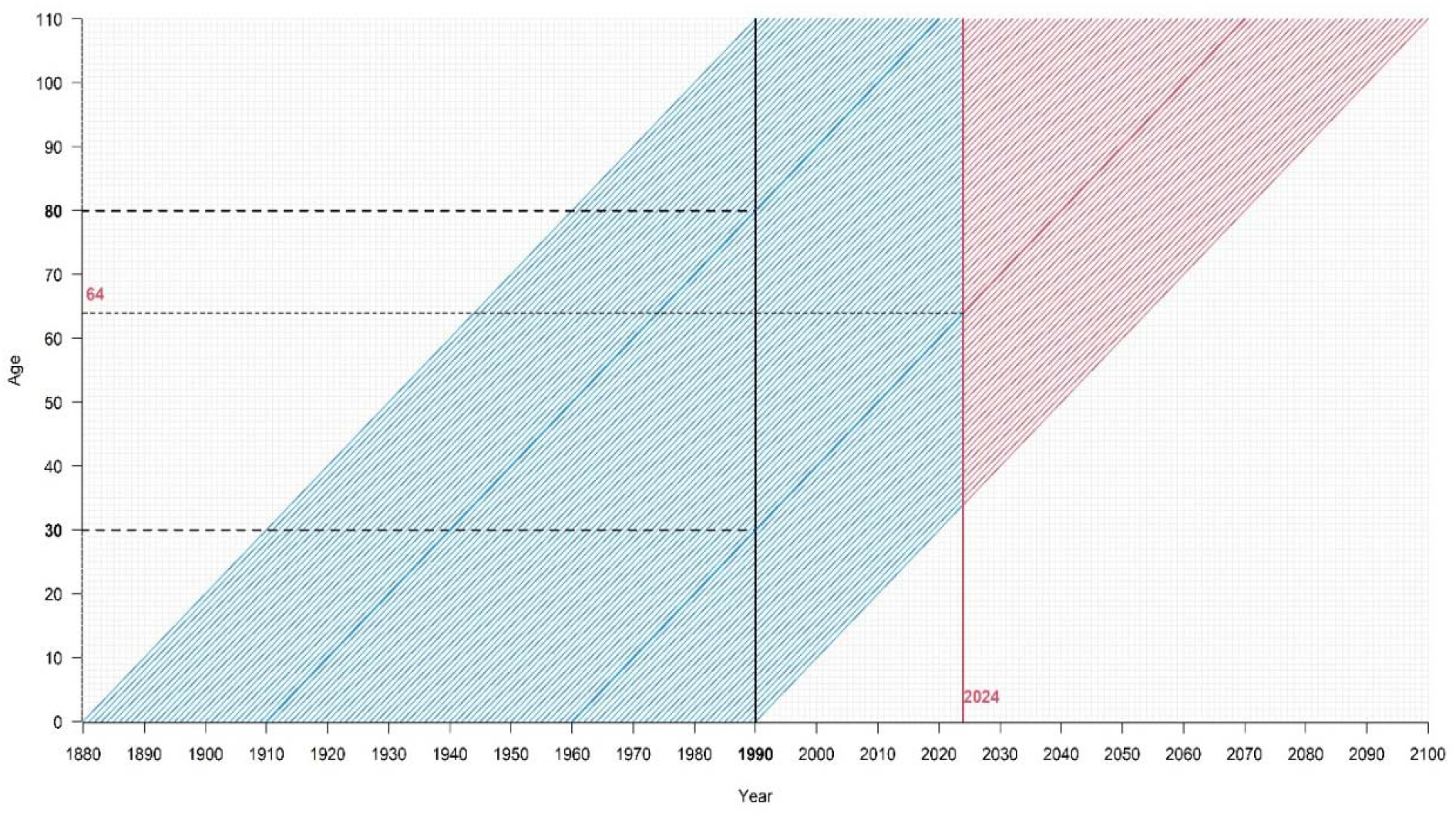
Lexis diagram of individual trajectories of active cohorts in 1990, some of which continue beyond the last year observed, 2024. As an illustration, the active cohort of individuals aged 80 in 1990 is fully observed, while mortality is not observed for the active cohort aged 30 in 1990 from age 64 onwards.

More precisely we adopted a log-linear Poisson regression model specified as follows:

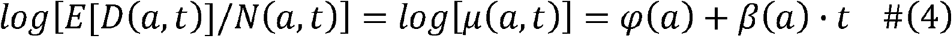

Here, *D* (*a,t*) is the number of deaths at age *a* in year *t*, the population size at the same age *N* (*a,t*) is considered as fixed in the model (*offset*), *φ* (*a*) is an age effect on log-mortality rates, and *β* (*a*) is an age-specific period effect, allowing a different piece of mortality change over time for each age (*interaction*). Both effects were modelled via natural splines with suitable choice of knots. The results of model (4) were plotted in the scale of the period life expectancy PLE, where we could verify goodness of fit and obtain PLE projections up to 2080, before being used to fill in the CLE of non-extinct cohorts (2) and obtain an estimate of the PoLE (1) over the period covered by the data.

Our results were also compared with those obtained with a simpler model assuming an age-independent mortality decline:

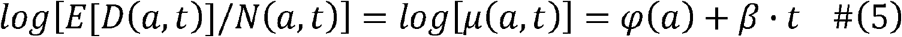

Note that model (5) was implicitly used by [15] to obtain their ACLE indicator, with *β* = log 1-*c*), where *C* was set to 0.5%.

## Results

The methods presented in the previous section have been applied to data for Switzerland and Norway from the Human Mortality Database (HMD). Data for these countries cover the periods 1876–2024 (Switzerland) and 1846–2024 (Norway). Figure 2 shows the period life expectancy PLE for both countries, separately for both sexes, between 1876 and 2024 (the earlier period for Norway is not shown due to high data variability).

**Figure 2.**
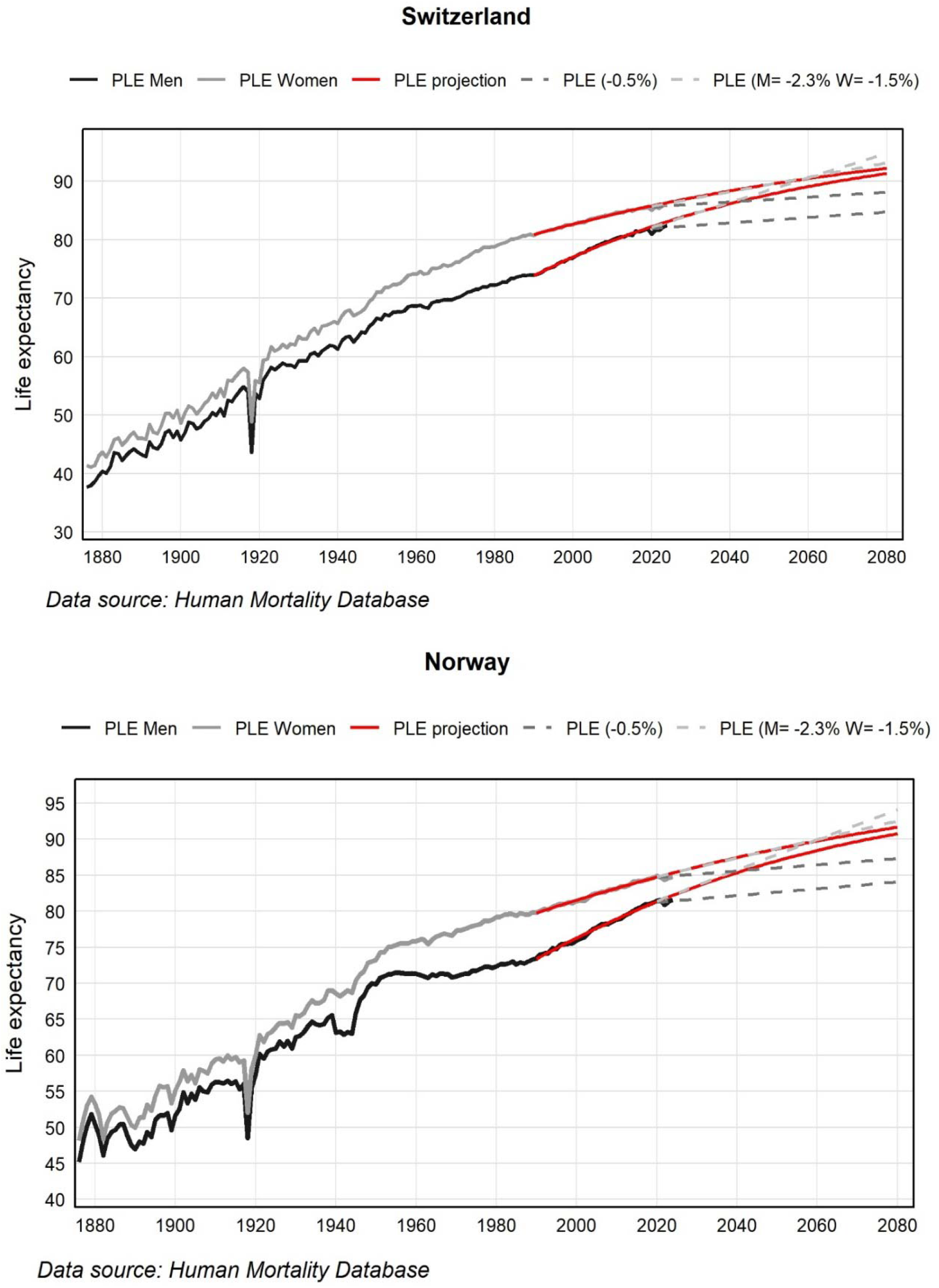
Period life expectancy (PLE) for Switzerland and Norway (1876-2024). Projections until 2080 were obtained using a model with age-dependent mortality decline (in red) or a model with age-independent mortality decline, either set at 5% (in dark gray) or estimated at 1.5% for women and 2.3% for men (in light gray).

Switzerland and Norway both experienced an impressive increase in PLE during this period. In Switzerland, PLE for women rose from 41.4 years in 1876 to 85.9 years in 2024, while for men it rose from 37.7 to 82.4 years. Women consistently had a higher PLE than men, but the gap between the two sexes has narrowed over time in the last three decades. The decline of nearly 10 years due to the Spanish flu in 1918 appears to have been the major demographic event of the last 150 years, while a less dramatic decline was also observed between 2020 and 2022 due to the COVID-19 pandemic, already described by [7-9]. In Norway, the increase in PLE between 1876 and 2024 was similarly marked, rising from 48.2 to 84.8 years for women, and from 45.2 to 81.6 years for men. Although PLE was more fluctuating in Norway than in Switzerland during the middle of the last century, the upward trend has been steady and similar in both countries over the past three decades, with a similar reduction in the gender gap.

The period between 1990 and 2019 was chosen as suitable for estimating the log-linear Poisson model (4) in both countries. On the one hand, a change in PLE trends was observed in the early 1990s (Figure 2) and, on the other hand, this choice excluded the outlier years marked by the COVID-19 pandemic. The results of the model are included in Figure 2, both the fit on the estimation period and projections up to 2080. The model fits the observed data well, while suggesting for the future a gradual slowdown in the PLE increase and a further narrowing of the gap between the two sexes.

Figure 2 also shows the PLE projections obtained assuming an age-independent annual decline in mortality, either of 0.5% (as in [15]) or estimated via model (5) on the period 1990-2019. Using model (5), a more important decline was estimated than the original 0.5%, namely a decline of 1.5% for women and 2.3% for men in both countries. In the scale of PLE, the 0.5% assumption significantly underestimates future gains (Figure 2), while using country and sex specific estimates of the age-independent mortality decline via model (5) PLE projections are more realistic over short periods, but they tend to increase too linearly and rapidly for men, with a crossing of the two sexes around 2060.

Mortality projections obtained with our model (4) were used to fill in the mortality of non-extinct cohorts in (3), providing for each year *y* and age *a*.the remaining cohort life expectancy CLE at age *a* of the cohort born in year *y* − *a*. Table 1 compares those remaining CLE with the remaining PLE for both countries and sexes and for selected years and ages. As a result of the mortality decline, CLE was systematically higher than PLE. In 2024, the difference at birth was 10.5 years for Swiss and Norwegian men, while it was of 7.7 and 8.3 years for Swiss and Norwegian women, respectively.

**Table 1:** Remaining period life expectancies (PLE), cohort life expectancies (CLE), and population life expectancies (PoLE) for Switzerland and Norway and both sexes and for selected years and ages.

|  |  | 1876 |  | 1900 |  | 1950 |  | 2000 |  | 2024 |  |
| --- | --- | --- | --- | --- | --- | --- | --- | --- | --- | --- | --- |
|  |  | Swiss | Norway | Swiss | Norway | Swiss | Norway | Swiss | Norway | Swiss | Norway |
| MEN | Age | PLE/CLE | PLE/CLE | PLE/CLE | PLE/CLE | PLE/CLE | PLE/CLE | PLE/CLE | PLE/CLE | PLE/CLE | PLE/CL<br>E |
|  | 0 | 37.7/41.7 | 45.2/49.6 | 45.8/52.5 | 51.7/56.4 | 66.6/77.0 | 69.9/77.2 | 76.9/90.0 | 76.0/89.4 | 82.4/92.9 | 81.6/92.1 |
|  | 30 | 30.5/32.1 | 35.9/38.6 | 33.0/36.0 | 37.6/39.5 | 41.2/44.8 | 44.3/44.5 | 48.2/56.5 | 47.3/55.7 | 53.1/60.2 | 52.3/59.6 |
|  | 50 | 17.3/18.4 | 22.0/23.4 | 18.5/19.0 | 23.2/24.1 | 23.4/24.4 | 26.1/25.5 | 29.5/33.8 | 28.6/33.1 | 33.9/38.1 | 33.2/37.6 |
|  | 65 | 9.2/9.7 | 12.4/12.8 | 9.9/10.0 | 12.9/13.6 | 12.6/12.9 | 14.4/14.6 | 16.9/19.0 | 16.1/18.1 | 20.4/22.6 | 19.8/22.0 |
|  | 80 | 4.0/4.3 | 5.7/5.7 | 4.2/4.3 | 5.3/5.7 | 5.4/5.2 | 6.0/6.3 | 7.3/7.5 | 6.8/7.1 | 9.4/9.7 | 8.8/9.2 |
|  | PoL<br>E | 63.3 | 66.7 | 66.7 | 69.9 | 76.8 | 77.0 | 86.2 | 85.7 | 89.7 | 89.1 |
| WOME<br>N | Age | PLE/CLE | PLE/CLE | PLE/CLE | PLE/CLE | PLE/CLE | PLE/CLE | PLE/CLE | PLE/CLE | PLE/CLE | PLE/CL<br>E |
|  | 0 | 41.4/46.2 | 48.2/53.3 | 48.6/58.8 | 55.1/62.2 | 71.1/82.9 | 73.2/82.4 | 82.6/91.5 | 81.4/90.8 | 85.9/93.6 | 84.8/93.1 |
|  | 30 | 32.6/34.8 | 37.3/39.9 | 34.7/38.8 | 39.3/41.2 | 44.5/51.3 | 46.4/51.0 | 53.4/59.1 | 52.1/58.2 | 56.4/61.6 | 55.3/60.9 |
|  | 50 | 18.6/19.7 | 23.5/24.7 | 19.5/21.1 | 24.6/25.2 | 26.2/29.4 | 27.8/30.2 | 34.1/37.3 | 32.9/36.2 | 36.9/40.1 | 35.7/39.3 |
|  | 65 | 9.5/9.7 | 13.3/13.8 | 9.8/10.6 | 13.8/14.5 | 14.3/15.2 | 15.4/16.2 | 20.7/22.3 | 19.7/21.3 | 23.0/24.8 | 21.9/24.0 |
|  | 80 | 4.2/4.3 | 6.1/6.1 | 4.3/4.6 | 5.8/6.1 | 5.9/5.8 | 6.4/6.7 | 9.1/9.3 | 8.6/9.1 | 10.8/11.2 | 10.2/10.8 |
|  | PoL<br>E | 65.4 | 69.0 | 69.9 | 71.9 | 82.0 | 81.9 | 88.9 | 88.2 | 91.3 | 90.7 |

At age 65, this difference was 2.2 years for Swiss and Norwegian men, while it was of 1.8 and 2.1 years for Swiss and Norwegian women, respectively.

Averaging the remaining CLE at each age via (1) we have then estimated our novel indicator, the population life expectancy PoLE. Results can be found in Table 1 for both sexes and countries and selected years. In Figure 3, the PoLE is compared with PLE and CLE over the entire period 1876-2024. PoLE significantly exceeded CLE and PLE in the early years. In 1900, the PLE, CLE, and PoLE for Swiss men were 45.8, 52.5, and 66.7, respectively. Similar differences were observed for both sexes and countries. Afterwards, PoLE showed a less sustained growth and a more regular trend than the other two measures. Around the mid of the 20th century, CLE overtook PoLE for both sexes and countries, although PoLE continued to consistently exceed PLE, including in most recent years. In 2024, the PoLE for Swiss men was 89.7 years, 3.2 years less than CLE (92.9 years), but 7.3 years more than PLE (82.4 years), these differences being of 2.3 and 5.4 years for Swiss women. Similar results were observed Norway (Table 1).

**Figure 3.**
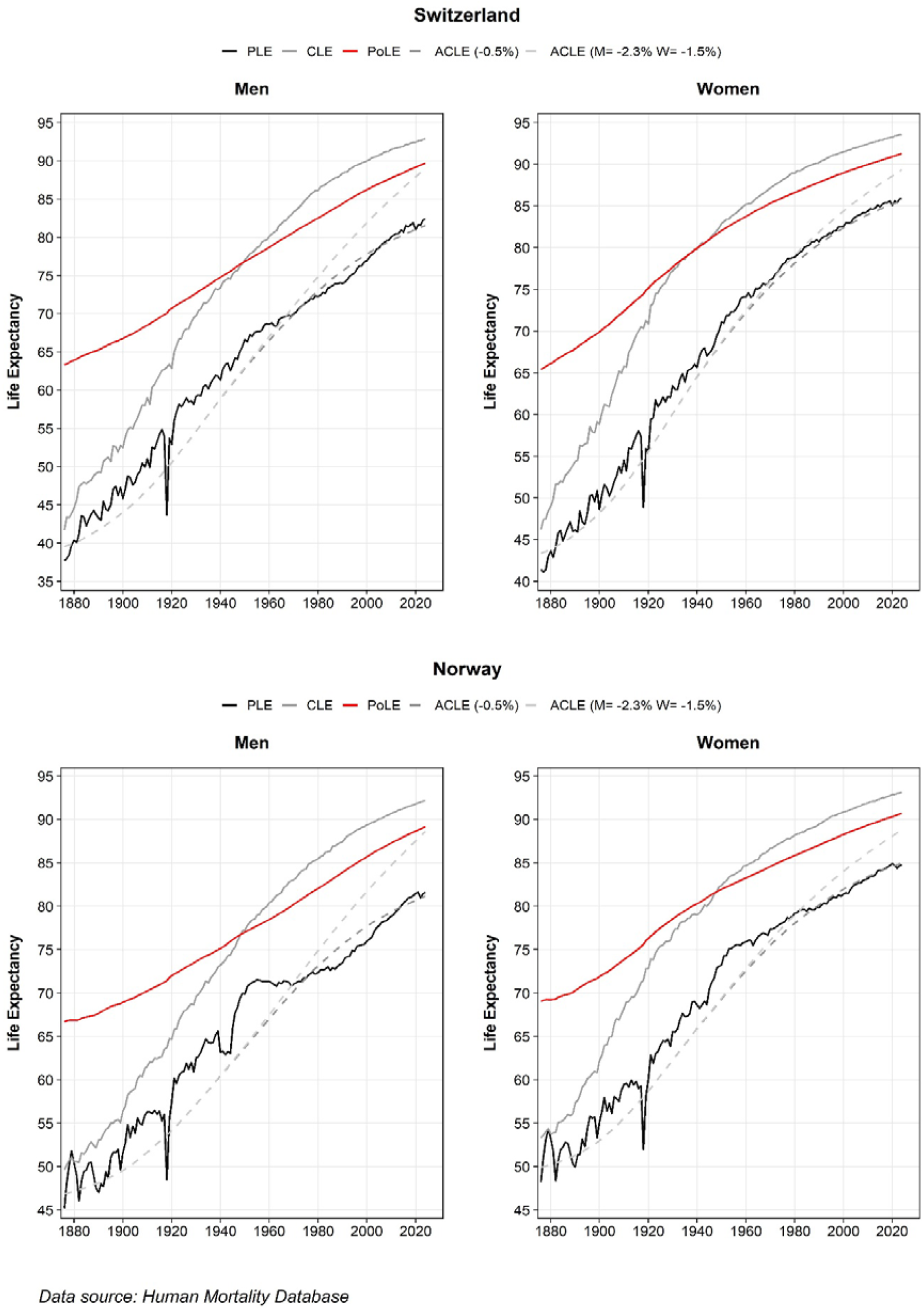
Period life expectancy (PLE), cohort life expectancy (CLE), population life expectancy (PoLE), and average cohort life expectancy (ACLE), the latter calculated with mortality decline either set at 0.5%, or estimated at 1.5% (women), or 2.3% (men). Switzerland and Norway (1876 - 2024).

Figure 3 also shows the average cohort life expectancy ACLE [15] on the same period, based on either the original assumption of a 0.5% annual mortality decline, or the (age-independent) mortality decline estimated via model (5), i.e. 1.5% for women and 2.3% for men in both countries. This indicator remained below the PLE for the period 1876-1980, regardless of the assumption. Using the 0.5% assumption, the ACLE slightly exceeded the PLE for the period 1980-2010 for men in both countries, while being almost equal to PLE for women. Using model (5), the ACLE was consistently above the PLE after 1980, while remaining significantly below the PoLE.

## Discussion

Period Life Expectancy (PLE) is a widely used indicator of longevity due to its many advantages, including its ability to respond to sudden crises or long-term societal changes [1]. However, this indicator remains difficult to interpret, as it refers to a hypothetical cohort, one that would live its entire life in a single year [1,10].

Following concerns and ideas of demographers over the past 40 years [11-20], we attempted in this paper to define a life expectancy indicator referring to a real population rather than a hypothetical cohort. A population is a mix of individuals of different ages, belonging to different cohorts that began living at different times in the past and will continue to live in the future. The first question that needs to be answered is therefore: how many years are people of a *given age*, i.e., belonging to a given “active cohort” of the population, likely to live in the future? To answer this question, which is important from both an individual and societal perspective, we need to make mortality projections based on past trends. Using a simple but well-fitting prediction model, we have notably estimated that a male newborn in 2024 can expect to live more than 10 years longer (and a female newborn more than 8 years longer) than suggested by the PLE at birth for the same year in Switzerland and Norway. Likewise, a 65-year-old living in 2024 can expect to live about 2 years longer than indicated by the remaining PLE at age 65.

Averaging such expected age at death over all ages, we could introduce a novel indicator that we have called Population Life Expectancy (PoLE), representing the mean age at death in a real population. The PoLE indicator has been calculated for Switzerland and Norway for the period 1876-2024 and was found to be systematically and considerably higher than the corresponding PLE. These results are clearly more realistic and convincing than those provided by other indicators that were supposed to measure the same concept, notably the Cross-sectional Average Length of life (CAL) [12] and the Average Cohort Life Expectancy (ACLE) [15], the former being systematically lower than PLE, the latter barely exceeding PLE (and only during certain periods), although it has been recognized that they should have been significantly higher than PLE in a context of declining mortality [13, 14]. We have detailed the differences between the definitions of our indicator and ACLE in the Methods section. The fundamental one is that ACLE is based on the life expectancy *at birth* of cohorts active in a given year, ignoring the fact that some years have *already* been lived by these cohorts and thereby underestimating their expected age at death, while PoLE is based on the *remaining* life expectancy of cohorts alive in a given year, thus properly estimating the expected age at death of the population. In other words, PoLE relies solely on the future mortality of active cohorts, while ACLE relies on both the future and past mortality of these cohorts (recall also that CAL was based only on past mortality, completely ignoring the future).

Although the PoLE indicator always exceeded PLE over the last 150 years, the former has increased at a slower pace than the latter, resulting in a more important difference during the first part of our observation period (reaching about 20 years in 1900), which gradually narrowed until recently (being still 5 to 8 years in 2024). The reason lies in the fact that infant mortality, which was high at the beginning of last century and then progressively declined due to improvement in medicine, hygiene and sanitary conditions [21-22], has a significant impact on PLE [23], while PoLE is only marginally affected. In fact, with the exception of newborns, all other cohorts active in a given year have survived the critical early life period and their chances of living longer are increased. Thus, at the beginning of last century, a man from the Swiss population could expect to live (and has lived!) up to more than 65 years, even though PLE for men at that time was only around 45, while a woman could expect to live up to 70 years, despite a PLE of less than 50. Our perception of the reality of mortality conditions for individuals living around 1900 is thus completely different using PoLE. For example, it is still worth remembering that a man who died at the age of 30 during World War I did not lose 15 years, but 35 years.

Comparing PoLE with the Cohort Life Expectancy (CLE) over the same period, an interesting phenomenon emerges. In the early 20th century, and until about the middle of the century, PoLE was also higher than CLE, which is strongly influenced by infant mortality (as is PLE), but the two curves crossed around 1950, CLE becoming higher than PoLE afterwards. Thus, before that year, life expectancy was greater for those who were alive than for those who were born (due to infant mortality), whereas after that year, life expectancy was greater for those who were born than for those who were alive (due to long term mortality decline). This crossing could thus be interpreted as a tangible sign of “human progress”, and it will certainly be worth to explore this further. A direction for future work will be to study the possible relation between the crossover of PoLE and CLE and the phases of the demographic transition [24]. This will be possible by extending the PoLE (and CLE) calculation to other countries with different demographic and health profiles, and specific historical trends in life expectancy.

In 2024, PoLE was estimated to be 2-3 years less than the CLE of a newborn, but it remains 5-8 years higher than PLE. Of course, these estimates involve some degree of uncertainty due to projections, which is a limitation of our approach, and somewhat different results would have been obtained using a different model, but, barring unforeseeable events, they at least give a fairly accurate idea of the relationships and rankings between these three concepts of life expectancy.

In this study, we adopted a projection model that only considers dimensions of time and age, with an interaction between the two, in order to describe mortality trends, which is rather classical [25]. The model fits the data well, and the resulting projections are credible in the long term, with a gradual slowdown in the increase in life expectancy and a narrowing (but not crossing) of the gender gap. We also showed the crucial importance of including the time-age interaction to account for different mortality trends at different ages. Without this, long-term projections of life expectancy would be unrealistic, overly linear with a crossover between the two sexes by 2060. However, additional work on the choice of the “best” projection model is still needed, exploring different options and variants of our model. For example, one could introduce a Gompertz assumption to describe mortality among older people [26], consider a quadratic effect of time on mortality to allow leveling off in the decline of mortality at some ages, and compare the fits obtained using different estimation periods. The last can in fact largely impact predictions, as shown for example by [27]. Finally, we would like to emphasize that the PoLE indicator proposed in this study is not intended to replace the classical PLE (or CLE), but to provide additional and complementary information about a population’s longevity. For example, PLE remains the best indicator for estimating the effect of a sudden event such as a pandemic, given its sensitivity to conjunctural changes in mortality. But PoLE will be a more accurate indicator to inform us on the actual gain in life expectancy achieved by a real population over a period of time. For example, PoLE shows that the gain in life expectancy of the (real!) Swiss and Norwegian population over 150 years corresponds to around +50% (from around 60 to 90 years) rather than +100% (from around 40 to 80 years), as PLE would suggest.

## Data Availability

All data in the present study are publicly available and can be accessed on the Human Mortality Database (HMD). All data produced in the present study are available in the manuscript.

## References

[1] M. Luy, P. D. Giulio, V. D. Lego, P. Lazarevič, et M. Sauerberg, « Life Expectancy: Frequently Used, but Hardly Understood », Gerontology, vol. 66, no 1, p. 95, août 2019, doi: 10.1159/000500955.

[2] M. Pechholdová, P. Grigoriev, F. Meslé, et J. Vallin, « Espérance de vie□: les deux Allemagne ont-elles convergé depuis la réunification□? »:, Popul. Sociétés, vol. N° 544, no 5, p. 1⍰4, mai 2017, doi: 10.3917/popsoc.544.0001.

[3] S. Harper, C. A. Riddell, et N. B. King, « Declining Life Expectancy in the United States: Missing the Trees for the Forest », Annu. Rev. Public Health, vol. 42, no 1, p. 381⍰403, avr. 2021, doi: 10.1146/annurev-publhealth-082619-104231.

[4] J. M. Aburto et al., « Quantifying impacts of the COVID-19 pandemic through life-expectancy losses: a population-level study of 29 countries », Int. J. Epidemiol., vol. 51, no 1, p. 63⍰74, sept. 2021, doi: 10.1093/ije/dyab207.

[5] G. Huang et al., « Changing impact of COVID-19 on life expectancy 2019–2023 and its decomposition: Findings from 27 countries », SSM - Popul. Health, vol. 25, p. 101568, déc. 2023, doi: 10.1016/j.ssmph.2023.101568.

[6] J. Schöley et al., « Life expectancy changes since COVID-19 », Nat. Hum. Behav., vol. 6, no 12, p. 1649⍰1659, éc. 2022, doi: 10.1038/s41562-022-01450-3.

[7] I. Locatelli et V. Rousson, « A first analysis of excess mortality in Switzerland in 2020 », PLoS ONE, vol. 16, no 6, p. e0253505, juin 2021, doi: 10.1371/journal.pone.0253505.

[8] I. Locatelli et V. Rousson, « Two complementary approaches to estimate an excess of mortality: The case of Switzerland 2022 », PLOS ONE, vol. 18, no 8, p. e0290160, août 2023, doi: 10.1371/journal.pone.0290160.

[9] I. Locatelli et V. Rousson, « Mortality in Switzerland in 2021 », PLoS ONE, vol. 17, no 9, p. e0274295, sept. 2022, doi: 10.1371/journal.pone.0274295.

[10] Goldstein, J., and R. Lee. 2020. “Demographic Perspectives on the Mortality of COVID-19 and Other Epidemics.” PNAS 117: 22035–22041.

[11] N. Brouard, « Structure et dynamique des populations. La pyramide des années à vivre, aspects nationaux et exemples régionaux », 1986, doi: 10.3406/espos.1986.1120.

[12] M. Guillot, « The Cross-Sectional Average Length of Life (CAL): A Cross-Sectional Mortality Measure That Reflects the Experience of Cohorts », Popul. Stud., vol. 57, no 1, p. 41⍰54, 2003.

[13] J. R. Goldstein, « Found in translation? A cohort perspective on tempo-adjusted life expectancy », in How Long Do We Live?, E. Barbi, J. W. Vaupel, et J. Bongaarts, Éd., in Demographic Research Monographs., Berlin, Heidelberg: Springer Berlin Heidelberg, 2008, p. 247⍰259. doi: 10.1007/978-3-540-78520-0_13.

[14] K. W. Wachter, « Tempo and its Tribulations », Demogr. Res., vol. 13, p. 201⍰222, nov. 2005, doi: 10.4054/DemRes.2005.13.9.

[15] R. Schoen et V. Canudas-Romo, « Changing mortality and average cohort life expectancy », Demogr. Res., vol. 13, p. 117⍰142, oct. 2005, doi: 10.4054/DemRes.2005.13.5.

[16] J. Bongaarts et G. Feeney, « Estimating mean lifetime », Proc. Natl. Acad. Sci. U. S. A., vol. 100, no 23, p. 13127⍰13133, nov. 2003, doi: 10.1073/pnas.2035060100.

[17] M. Guillot et H. S. Kim, « On the correspondence between CAL and lagged cohort life expectancy », Demogr. Res., vol. 24, p. 611⍰632, avr. 2011, doi: 10.4054/DemRes.2011.24.25.

[18] V. Canudas-Romo et M. Guillot, « Truncated cross-sectional average length of life: A measure for comparing the mortality history of cohorts », Popul. Stud., vol. 69, no 2, p. 147⍰159, mai 2015, doi: 10.1080/00324728.2015.1019955.

[19] M. R. Nepomuceno, Q. Cui, A. van Raalte, J. M. Aburto, et V. Canudas-Romo, « The Cross-sectional Average Inequality in Lifespan (CAL†): A Lifespan Variation Measure That Reflects the Mortality Histories of Cohorts », Demography, vol. 59, no 1, p. 187⍰206, févr. 2022, doi: 10.1215/00703370-9637380.

[20] R. Mogi, J. Nisén, et V. Canudas-Romo, « Cross-Sectional Average Length of Life Childless », Demography, vol. 58, no 1, p. 321⍰344, févr. 2021, doi: 10.1215/00703370-8937427.

[21] J. Vallin et F. Meslé, « Convergences and divergences in mortality: A new approach of health transition », Demogr. Res., vol. S2, p. 11⍰44, avr. 2004, doi: 10.4054/DemRes.2004.S2.2.

[22] D. Cutler et G. Miller, « The role of public health improvements in health advances: the twentieth-century United States », Demography, vol. 42, no 1, p. 1⍰22, févr. 2005, doi: 10.1353/dem.2005.0002.

[23] J. Véron, S. Preston, P. Heuveline, et M. Guillot, « Demography, Measuring and Modeling Population Processes », Popul. Fr. Ed., vol. 57, p. 591, mai 2002, doi: 10.2307/1535065.

[24] A. R. Omran, « The epidemiologic transition. A theory of the Epidemiology of population change. 1971. », Bull. World Health Organ., vol. 79, no 2, p. 161⍰170, 2001.

[25] R. D. Lee et L. R. Carter, « Modeling and Forecasting U. S. Mortality », J. Am. Stat. Assoc., vol. 87, no 419, p. 659⍰671, 1992, doi: 10.2307/2290201.

[26] Gavrilov LA, Gavrilova NS (2019) New Trend in Old-Age Mortality: Gompertzialization of Mortality Trajectory. Gerontology, 65(5):451–457. doi: 10.1159/000500141.

[27] T. Ferenci (2023) Comparing methods to predict baseline mortality for excess mortality calculations. BMC Med Res Methodol 23(1):239. doi: 10.1186/s12874-023-02061-w.

